# Immunogenicity of a third dose of BNT162b2 to ancestral SARS-CoV-2 & Omicron variant in adults who received two doses of inactivated vaccine

**DOI:** 10.1101/2022.01.20.22269586

**Authors:** Nancy H. L. Leung, Samuel M. S. Cheng, Mario Martín-Sánchez, Niki Y. M. Au, Yvonne Y. Ng, Leo L. H. Luk, Karl C. K. Chan, John K. C. Li, Yonna W. Y. Leung, Leo C. H. Tsang, Sara Chaothai, Kelvin K. H. Kwan, Dennis K. M. Ip, Leo L. M. Poon, Gabriel M. Leung, J. S. Malik Peiris, Benjamin J. Cowling

## Abstract

**Background:** Limited data exist on antibody responses to mixed vaccination strategies involving inactivated COVID-19 vaccines, particularly in the context of emerging variants.

**Methods:** We conducted an open label trial of a third vaccine dose of an mRNA vaccine (BNT162b2, Fosun Pharma/BioNTech) in adults aged ≥30 years who had previously received two doses of inactivated COVID-19 vaccine. We collected blood samples before administering the third dose and 28 days later, and tested for antibodies to the ancestral virus using a binding assay (ELISA), a surrogate virus neutralization test (sVNT) and a live virus plaque reduction neutralization test (PRNT). We also tested for antibodies against the Omicron variant using live-virus PRNT.

**Results:** In 315 participants, a third dose of BNT162b2 substantially increased antibody titers on each assay. Mean ELISA levels increased from an optical density (OD) of 0.3 to 2.2 (p<0. 001), and mean sVNT levels increased from an inhibition of 17% to 96% (p<0.001). In a random subset of 20 participants, the geometric mean PRNT_50_ titers rose very substantially by at least 24 fold from Day 0 to Day 28 against the ancestral virus (p<0.001) and rose by at least 11 fold against the Omicron variant (p<0.001). In daily monitoring, post-vaccination reactions subsided within 7 days for over 99% of participants.

**Conclusions:** A third dose of COVID-19 vaccination with an mRNA vaccine substantially improved antibody levels against the ancestral virus and the Omicron variant with well-tolerated safety profile, in adults who had received two doses of inactivated vaccine 6 months earlier.

**Summary:** In this open label trial of Chinese adults aged ≥30 years who received two doses of inactivated COVID-19 vaccine 6 months earlier, third-dose mRNA vaccine substantially improved antibody levels against the ancestral virus and Omicron variant with well-tolerated safety profile.

## INTRODUCTION

The accrual of population immunity to COVID-19, acquired through natural infection or vaccination, will eventually bring an end to the pandemic and allow life to return to normal. Vaccine technologies being used for COVID-19 include inactivated virus, viral vectored and mRNA vaccines [1]. We have previously shown that two doses of the mRNA vaccine BNT162b2 (BioNTech/Fosun Pharma) conferred approximately 10-fold higher post-vaccination neutralising antibody titers than two doses of the aluminium hydroxide-adjuvanted inactivated virus vaccine CoronaVac (Sinovac) [2,3], while T cell responses to the two vaccines were similar [3]. The emergence of variants of concern (VOCs) and decreases in vaccine effectiveness within a few months after the second vaccine dose have resulted in recommendations for third doses [4].

Various studies have reported immunogenicity and safety of third doses using the same vaccine, i.e. homologous booster vaccination [5,6], but there are few studies evaluating heterologous booster, such as the use of a third dose of an mRNA vaccine in individuals who previously received two doses of inactivated vaccine [7-11]. Costa Clemens et al. conducted a non-inferiority randomised trial of a heterologous third dose of BNT162b2 or other vaccines against a homologous third dose of CoronaVac in adults who had received two doses of CoronaVac [9]. They showed that all heterologous booster groups have substantial rise in neutralizing antibody titers (FRNT_50_) against the Omicron and Delta variants [9]. They also reported more local reactions but less systemic reactions to BNT162b2 compared to two adenoviral vectored vaccines given as third doses. Pérez-Then et al. reported a third dose of BNT162b2 vaccination conferred a 1.4-fold increase in neutralizing antibodies (PRNT_50_) against Omicron when compared to two doses of an mRNA vaccine (BNT162b2 or mRNA-1273) [8]. Here, we report a trial of the immunogenicity and reactogenicity to a third dose of BNT162b2 in Chinese adults who had previously received two doses of inactivated COVID-19 vaccine.

## MATERIALS AND METHODS

### Study design

The “<u>m</u>RNA vaccination to <u>boost</u> antibodies against SARS-CoV-2 in recipients of inactivated vaccines” (mBoost) study is an open-label single-arm clinical trial to measure the antibody responses and reactogenicity of an mRNA vaccine (BNT162b2) given as a third dose in adults ≥30 years of age who have previously received two doses of an inactivated COVID-19 vaccine. CoronaVac and BNT162b2 have been available to adults in Hong Kong since March 2021. Some Hong Kong residents could have received two doses of inactivated vaccine BIBP (Sinopharm) instead from mainland China or overseas. The BNT162b2 vaccine used in Hong Kong, known as the Pfizer/BioNTech vaccine elsewhere, are distributed solely by Fosun Pharma in Greater China.

Enrolment invitations were extended to community-dwelling adults in Hong Kong through mass promotion efforts including advertisements in newspapers and social media. Interested adults were invited to visit the study website and complete an online screening form for initial assessment of enrolment eligibility, and confirmed again shortly before vaccination. Individuals were eligible if they were aged ≥30 years, had previously received two doses of an inactivated COVID-19 vaccine with the most recent dose ≥90 days prior to enrolment. We excluded individuals who reported a history of COVID-19 infection, received any dose of COVID-19 vaccine other than an inactivated vaccine, or who were unsuitable to receive an mRNA vaccine including but not limited to allergies to the active substance or other ingredients of the vaccine. We also excluded individuals with diagnosed medical conditions related to their immune system, use of medication that impairs immune system in the last 6 months except topical steroids or short-term oral steroids (course lasting ≤14 days), those who had used immunoglobulins or any blood products within 90 days prior to enrolment, and any females who were pregnant or intending to become pregnant in the coming 3 months.

We collected 20ml clotted blood specimens at the day of enrolment and vaccination (day 0) and again after 28 days, with additional blood draws planned after 182 and 365 days. This was a single-arm study with no need for randomization, and the participants and the study staff were aware of the type of vaccination administered (no blinding). After vaccination, participants were observed for 30 minutes to record any immediate events. We then asked participants to report the presence of a list of (post-vaccination) local or systematic events/reactions daily for 7 days using an online e-diary. If the participant was still reporting any events/reaction on Day 7, additional daily monitoring continued for up to three additional weeks until symptoms resolved. On each day, participants were also asked to grade whether the reported symptoms overall have interfered with their usual activities (mild, moderate and severe). Participants are interviewed at 28, 182 and 365 days after vaccination to collect information on underlying medical conditions, and any medical encounters including hospitalizations during the study. Participants were provided with a free tympanic thermometer at enrolment, and a gift voucher of HK$100 (US$13) at the follow-up blood draws. Study data were collected and managed using REDCap electronic data capture tools [12,13].

### Ethics

Written informed consent was obtained from all participants. The study protocol was approved by the Institutional Review Board of the University of Hong Kong. The study was registered on Clinicaltrials.gov prior to commencement (NCT05057182).

### Outcome measures

The primary outcome measure is the vaccine immunogenicity at 28 days after receipt of the third dose of BNT162b2, measured as geometric mean titer (GMT) of SARS-CoV-2 serum neutralizing antibodies against the vaccine strain (i.e. the ancestral virus) using plaque reduction neutralization test (PRNT). As the Omicron strain emerged in late 2021, we also evaluated vaccine immunogenicity against the Omicron strain. The secondary outcome measures include the GMT at other post-vaccination timepoints (i.e. at days 182 and 365) and mean-fold rises in antibody titers from baseline to each post-vaccination timepoint, and the incidence of solicited reactions or medical encounters following vaccination.

### Sample size justification

For the primary outcome of vaccine immunogenicity, i.e. the GMT of SARS-CoV-2 serum neutralizing antibodies against the vaccine strain (ancestral virus) at 28 days after vaccination, a sample size of 300 individuals was chosen based of feasibility [14].

### Laboratory methods

Blood samples were delivered to our study laboratory at the University of Hong Kong as soon as possible, with the optimal delivery time within 24h. Sera were extracted from the clotted blood within 48h and stored at -80°C until subsequent testing. We tested sera heat inactivated at 56°C for 30 minutes with three assays, our in-house enzyme-linked immunosorbent assay (ELISA) for the receptor binding domain (RBD) of the spike protein, a surrogate virus neutralisation test (sVNT) (GenScript), and a plaque reduction neutralisation test (PRNT) as previously described [11,15,16]. We aimed to tested sera collected at baseline and Day 28 with the ELISA and sVNT in all participants, and with PRNT in a randomly-selected subset of 20 participants. We have demonstrated a good correlation (r=0.77) between PRNT_50_ and sVNT neutralization percentage for ancestral virus in our earlier studies [17]. The ELISA was not designed as a quantitative assay, although it has a dynamic range of between 0 to 5 and we have demonstrated a good correlation (r=0.83) between ELISA OD_450_ and sVNT neutralization percentage for ancestral virus [17]. For ELISA a single 1:100 and for sVNT a single 1:10 serum dilution was tested respectively. For PRNT, a serial two-fold serum dilutions from 1:10 to 1:320 were tested. PRNT assays were carried out using ancestral SARS-CoV-2 BetaCoV/Hong Kong/VM20001061/2020 isolated in Hong Kong in January 2020 in Vero-E6 cells (ATCC CRL-1586), the Pango lineage B.1.1.529 Omicron variant designated hCoV-19/Hong Kong/VM21044713_WHP5047-S5/2021 isolated in Vero-E6 TMRSS2 cells (Vero E6 cells overexpressing TMPRSS2, kindly provided by Dr S Matsuyama and colleagues), and the passage level 3 virus aliquots were used. Cells were maintained in DMEM medium supplemented with 10% FBS and 100 U/ml penicillin– streptomycin (all from ThermoFisher Scientific, Waltham, MA, USA). Virus sequences are available in GISAID as EPI_ISL_412028 and EPI_ISL_6716902. The PRNT_50_ and PRNT_90_ titers were the highest serum dilutions neutralizing ≥50% and ≥90% of input plaques, respectively. The WHO control serum provided by the National Institute for Biological Standards and Control 20/136 gave PRNT_50_ titers of 320 and 320 against the ancestral virus and 20 and 40 against the Omicron variant in two titrations [11].

### Statistical analysis

We assessed the GMT of SARS-CoV-2 (PRNT) neutralizing antibody titers, surrogate virus neutralization percentages and the mean concentrations of SARS-CoV-2 Spike RBD IgG (proxy by OD_450_) at Day 28. For sVNT, measured negative neutralization percentages were transformed to zero. PRNT titers were taken as the reciprocal of the serum dilution and were interval-censored, e.g. a sample that was able to neutralise virus at a 1:20 dilution but not at a 1:40 dilution was reported as 20 to indicate that the titer was ≥20 and <40. We imputed titers <10 with the value 5 and titers ≥320 with the value 640 for estimation of GMTs. We compared antibody levels at Day 28 after receipt of BNT162b2 to baseline using Wilcoxon signed rank tests. Correlation of PRNT_90_ titers measured at Day 28 against the ancestral strain and the Omicron variant was estimated by Spearman’s rank correlation coefficient. Reactogenicity endpoints were described as frequency (%) for local and systemic reactions or events among participants who reported health status for at least one day in the week following receipt of the third dose of BNT162b2. Statistical analyses were conducted using R version 4.1.2.

## RESULTS

From 13 October 2021 through 18 January 2022 we screened 436 individuals, of which 366 (84%) were eligible and 315 (86%) were enrolled and administered BNT162b2 vaccination. We collected paired Day 0 and Day 28 blood samples in 312 (99%) vaccinated participants. Most of the participants were older Chinese adults (median aged 54 years, IQR 47-62), with 35% who were obese, and about one-third reported at least one chronic medical condition including hypertension (18%), hypercholesterolemia (13%) and diabetes (7%), and almost all (98%) reported receiving two doses of CoronaVac rather than other inactivated vaccines (Table 1). Although adults who had received two doses of inactivated virus vaccination at least 90 days ago were eligible to enrol into our study (Figure S1), 75% of participants reported receiving the second dose typically around 6-7 months earlier (Table 1). A detailed flow chart of participant enrolment is provided in Figure S1. The study is ongoing and Day 182 and 365 samples will be collected in Spring 2022 and Autumn 2022 respectively.

**Table 1.**
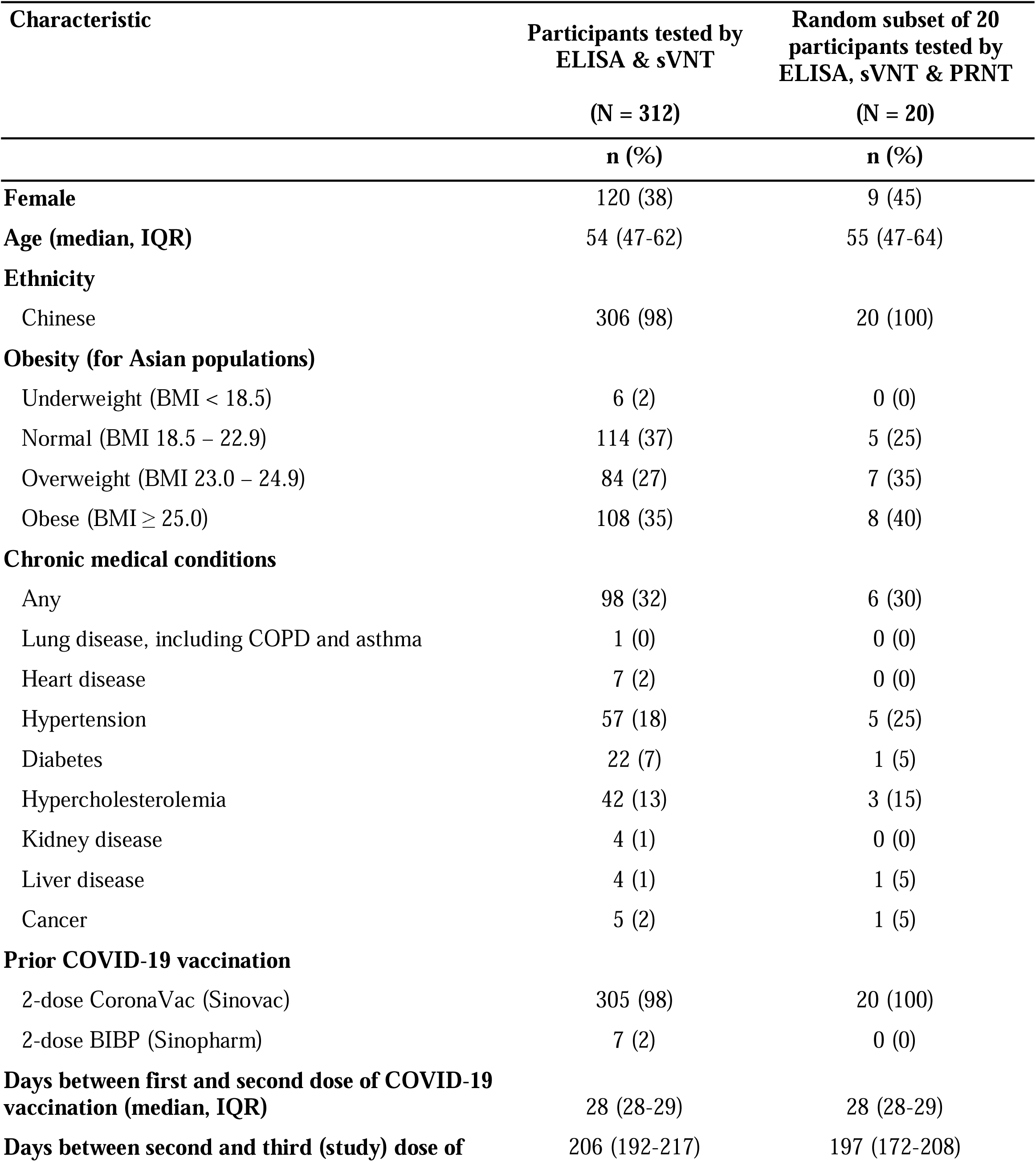
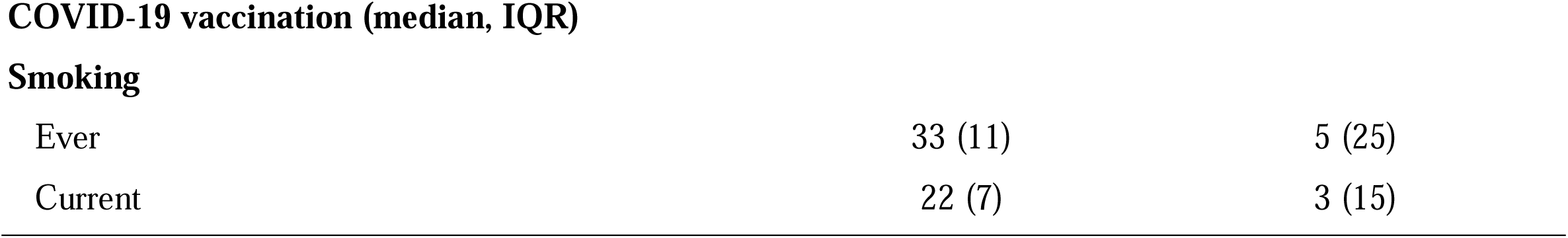
Characteristics of study participants at baseline. We enrolled and administered BNT162b2 vaccine as a third vaccine dose in 315 adults who previously received two doses of inactivated COVID-19 vaccine. Among them, ELISA and sVNT against ancestral SARS-CoV-2 virus were performed in paired Day 0 and Day 28 sera available from 312 vaccinated participants, and we randomly selected 20 participants to also perform PRNT against ancestral SARS-CoV-2 virus and the Omicron variant. ELISA: enzyme-linked immunosorbent assay; sVNT: surrogate virus neutralisation test; PRNT: plaque reduction neutralisation test.

| Characteristic | Participants tested by<br>ELISA & sVNT<br>(N = 312) | Random subset of 20<br>participants tested by<br>ELISA, sVNT & PRNT<br>(N = 20) |
| --- | --- | --- |
|  | n (%) | n (%) |
| **Female** | 120 (38) | 9 (45) |
| **Age (median, IQR)** | 54 (47-62) | 55 (47-64) |
| **Ethnicity** |  |  |
| Chinese | 306 (98) | 20 (100) |
| **Obesity (for Asian populations)** |  |  |
| Underweight (BMI < 18.5) | 6 (2) | 0 (0) |
| Normal (BMI 18.5 – 22.9) | 114 (37) | 5 (25) |
| Overweight (BMI 23.0 – 24.9) | 84 (27) | 7 (35) |
| Obese (BMI ≥ 25.0) | 108 (35) | 8 (40) |
| **Chronic medical conditions** |  |  |
| Any | 98 (32) | 6 (30) |
| Lung disease, including COPD and asthma | 1 (0) | 0 (0) |
| Heart disease | 7 (2) | 0 (0) |
| Hypertension | 57 (18) | 5 (25) |
| Diabetes | 22 (7) | 1 (5) |
| Hypercholesterolemia | 42 (13) | 3 (15) |
| Kidney disease | 4 (1) | 0 (0) |
| Liver disease | 4 (1) | 1 (5) |
| Cancer | 5 (2) | 1 (5) |
| **Prior COVID-19 vaccination** |  |  |
| 2-dose CoronaVac (Sinovac) | 305 (98) | 20 (100) |
| 2-dose BIBP (Sinopharm) | 7 (2) | 0 (0) |
| **Days between first and second dose of COVID-19**<br>**vaccination (median, IQR)** | 28 (28-29) | 28 (28-29) |
| **Days between second and third (study) dose of** | 206 (192-217) | 197 (172-208) |
| **Smoking** |  |  |
| Ever | 33 (11) | 5 (25) |
| Current | 22 (7) | 3 (15) |

The third dose of BNT162b2 substantially increased antibody titers measured by the various assays (Figure 1). Mean ELISA levels increased from a OD of 0.3 to 2.2 (p<0.001), and mean sVNT levels increased from an inhibition of 17% to 96% (p<0.001) (Table 2). We randomly selected a subset of 20 participants for further PRNT testing against the ancestral virus and the Omicron strain (Table 1). The ELISA and sVNT levels in these 20 participants were similar to that of the 312 vaccinated participants (Figure S2). In these 20 participants, the geometric mean PRNT_50_ titer against the ancestral strain at Day 0 was 12. We could not estimate the GMT at Day 28 because 18/20 (90%) samples were positive at the highest dilution of 1:320, and we noted that if the GMT were ≥320 it would correspond to a mean-fold titer rise of ≥24 from Day 0 to Day 28 (p<0.001). The geometric mean PRNT_90_ titer against the ancestral strain at Day 0 was 6. At Day 28, 12/20 samples were positive at the highest dilution of 1:320, and we noted that if the GMT were ≥320 it would correspond to a mean-fold titer rise of ≥35 in PRNT_90_ titers from Day 0 to Day 28 (p<0.001). Furthermore, in these same 20 participants the geometric mean PRNT_50_ titer against the Omicron strain at Day 0 was 5, and rose to 59 at Day 28, a mean-fold rise of at least 11 given that the Day 0 titers were almost all assigned values of 5 corresponding to the floor of the assay at <10 (p<0.001). The geometric mean PRNT_90_ titer against the Omicron strain at Day 0 was also 5, and rose to 19 at Day 28, a mean-fold rise of at least 3 (p<0.001). We did not identify any evidence that suggest a correlation between the PRNT_90_ titers against the Omicron variant and that against the ancestral virus (r=0.30, p=0.21) (Figure S3).

**Table 2.**
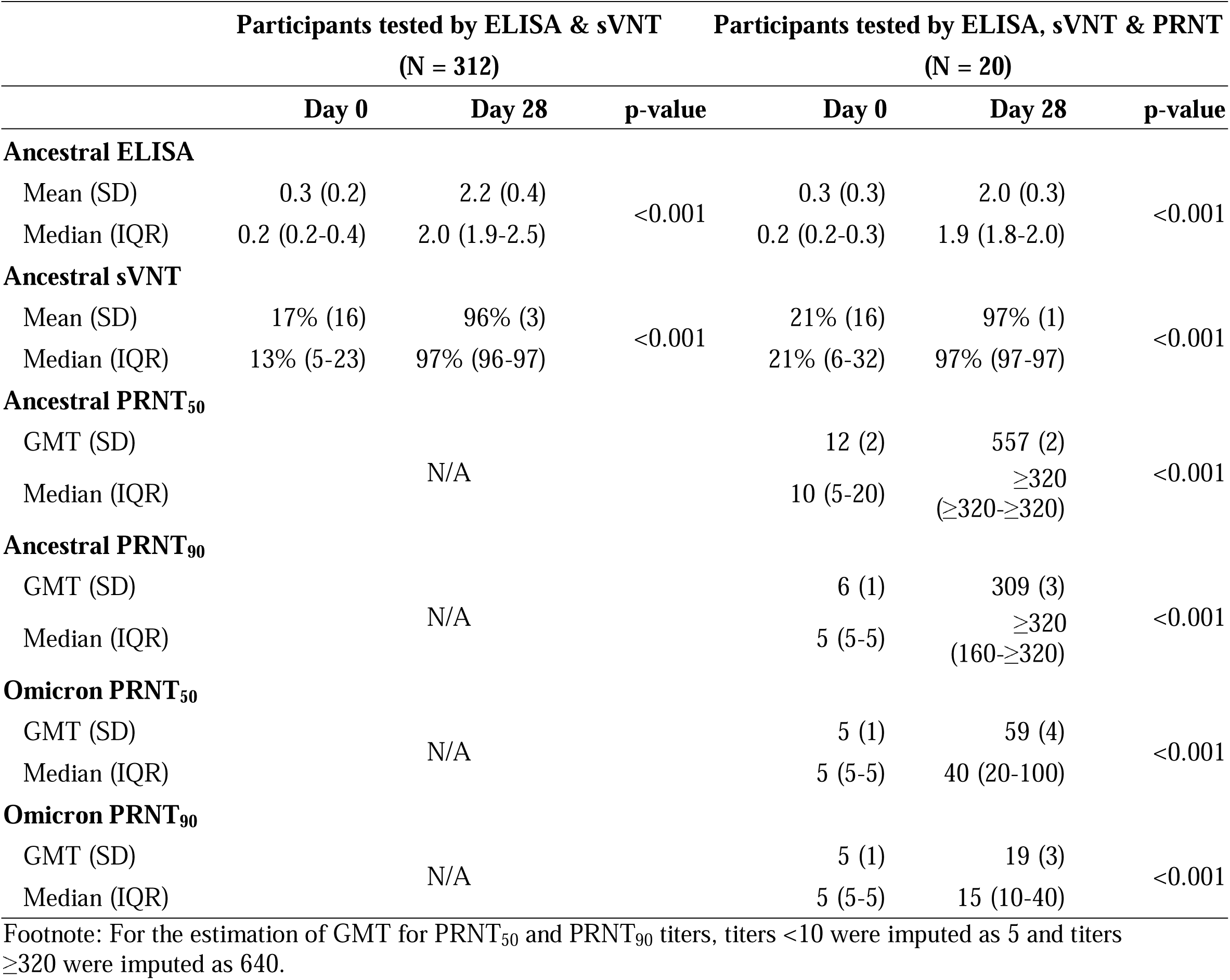
Antibody response after third dose BNT162b2 vaccination in adults who previously received two doses of inactivated COVID-19 vaccine. ELISA and sVNT against ancestral SARS-CoV-2 virus were performed in paired Day 0 and Day 28 sera available from 312 vaccinated participants, and we randomly selected 20 participants to also perform PRNT against ancestral SARS-CoV-2 virus and the Omicron variant. Significant differences between antibody level at Day 0 and Day 28 was evaluated by Wilcoxon signed rank tests. ELISA: enzyme-linked immunosorbent assay; sVNT: surrogate virus neutralisation test; PRNT: plaque reduction neutralisation test; GMT: geometric mean titer.

**Figure 1.**
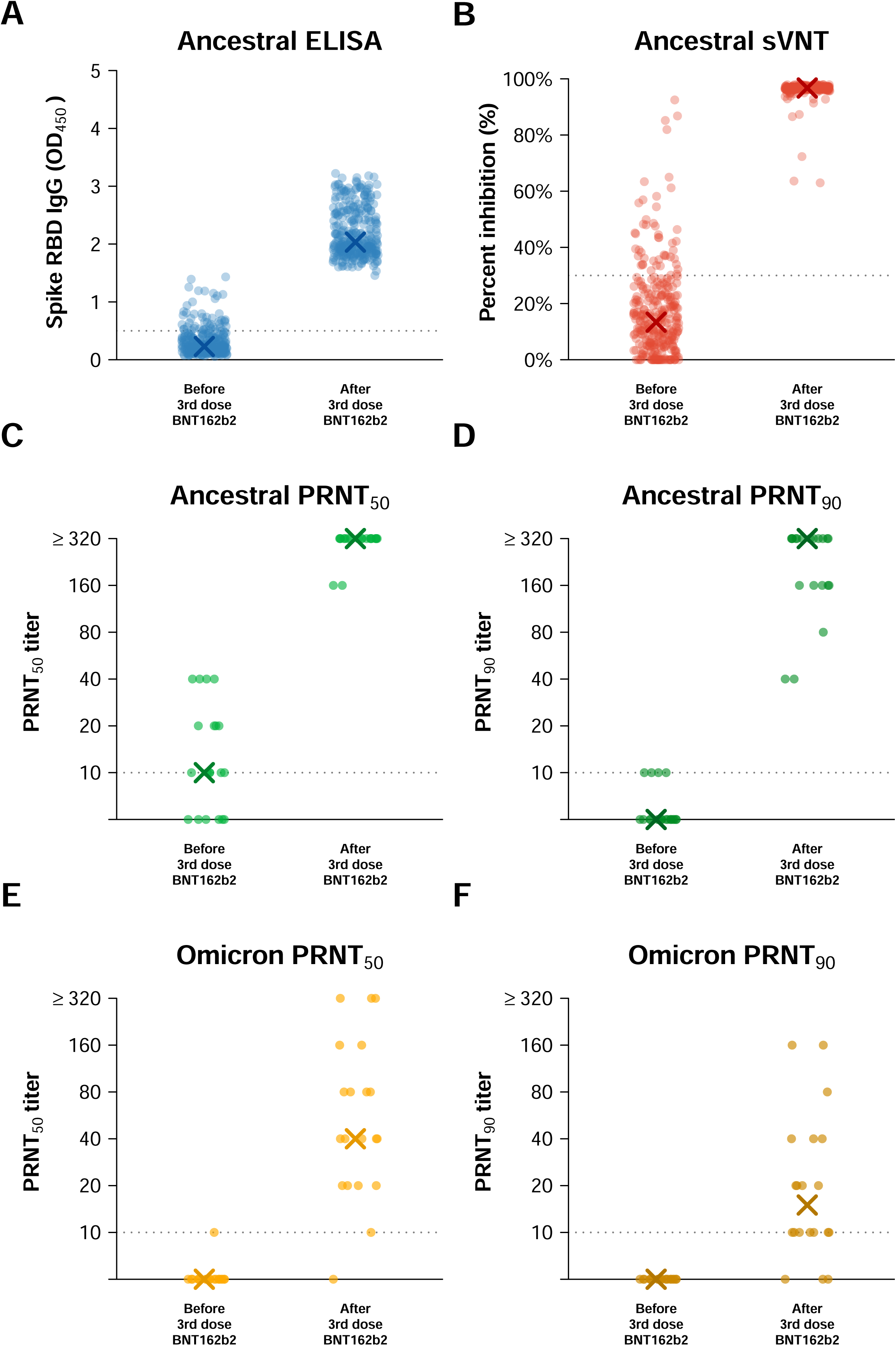
Third dose of SARS-CoV-2 BNT162b2 vaccine boosts serum antibodies against ancestral strain and Omicron variant in adults who previously received two doses of inactivated vaccines. **(A)** ELISA for serum IgG against SARS-CoV-2 Spike receptor binding domain (RBD) of ancestral strain. **(B)** Surrogate virus neutralization test (sVNT) against ancestral strain. Live virus plaque reduction neutralization test (PRNT) against ancestral strain with endpoints at **(C)** 50% (PRNT_50_) or **(D)** 90% (PRNT_90_). Live virus plaque reduction neutralization test (PRNT) against Omicron variant with endpoints at **(E)** 50% (PRNT_50_) or **(F)** 90% (PRNT_90_). X in each panel indicates the median level. Data for ELISA and sVNT were available from 312 vaccinated participants whom paired day 0 and day 28 sera were collected, while data for PRNT were available from a random sample of 20 participants.

Among the 315 participants who received BNT162b2 vaccination, 304 (97%) participants reported health status for at least one day in the week following receipt of BNT162b2, including 234 (77%) who reported every day and another 38 (13%) reported for ≥7 days. 193/304 (63%) participants reported feeling unwell for an average of 2.9 days (SD 1.8 days), and only 3 (<1%) participants reported feeling unwell beyond 7 days after third-dose BNT162b2 vaccination. Within the 7 days after receipt of BNT162b2, the most commonly reported local reactions were pain (46%) and tenderness (44%) at the injection site, while systemic reactions were reported by a minority of participants (Table 3). These symptoms usually subsided within 7 days after vaccination (Figure 2). Among 312 participants whose information is available, 6 (2%) reported having sought medical consultation within one month of the third dose but none were hospitalized. Among these 6 participants, 4 reported seeking medical consultation possibly for discomfort associated with vaccination within the week after vaccination, and the remaining two participants sought medical consultation due to back pain or stress outside the 7-day window.

**Table 3.** Solicited local and systemic reactions on the day after and anytime within 7 days after third dose BNT162b2 vaccination in adults who previously received two doses of inactivated COVID-19 vaccine. Daily frequencies within 7 days after third dose BNT162b2 vaccination for reactions highlighted in bold are shown in Figure 2.

| Post-vaccination reactions | Anytime within 7 days after third dose BNT162b2 vaccination<br>(N = 304)<br>n (%) | Day 1 after third dose BNT162b2 vaccination<br>(N = 304)<br>n (%) |
| --- | --- | --- |
| **<u>Local reactions at the injection site</u>** |  |  |
| **Pain\*** | **141 (46)** | **130 (43)** |
| **Tenderness\*** | **133 (44)** | **108 (36)** |
| **Swelling or hardness\*** | **71 (23)** | **51 (17)** |
| **Itchiness\*** | **20 (7)** | **2 (1)** |
| **Redness\*** | **15 (5)** | **5 (2)** |
| **<u>Systemic reactions and general symptoms</u>** |  |  |
| **Feverish** | **72 (24)** | **55 (18)** |
| **Fever $\geq 38.0^{\circ}\text{C}$** | **26 (9)** | **20 (7)** |
| **Chills** | **32 (11)** | **29 (10)** |
| **Headache** | **75 (25)** | **56 (18)** |
| **Myalgia** | **54 (18)** | **38 (12)** |
| **Fatigue** | **56 (18)** | **42 (14)** |
| **Drowsiness** | **51 (17)** | **38 (12)** |
| **Malaise** | **31 (10)** | **20 (7)** |
| Pain at the injection arm | 25 (8) | 14 (5) |
| Loss of appetite | 21 (7) | 12 (4) |
| Dizziness | 21 (7) | 11 (4) |
| Runny nose | 20 (7) | 12 (4) |
| Nasal congestion | 19 (6) | 9 (3) |
| Arthralgia | 17 (6) | 10 (3) |
| Sneezing | 17 (6) | 8 (3) |
| Sore throat | 17 (6) | 10 (3) |
| Chest discomfort | 13 (4) | 9 (3) |
| Diarrhoea | 13 (4) | 4 (1) |
| Abdominal distention | 11 (4) | 6 (2) |
| Palpitations | 11 (4) | 5 (2) |
| Insomnia | 11 (4) | 5 (2) |
| Phlegm | 10 (3) | 5 (2) |
| Nausea | 9 (3) | 5 (2) |
| Cough | 8 (3) | 4 (1) |
| Abdominal pain | 7 (2) | 4 (1) |
| Constipation | 6 (2) | 4 (1) |
| Body itching | 5 (2) | 2 (1) |
| Flushing of the face | 4 (1) | 3 (1) |
| Skin rash | 3 (1) | 1 (0) |
| Chest pain | 3 (1) | 0 (0) |
| Enlarged lymph nodes | 3 (1) | 0 (0) |
| Face swelling | 2 (1) | 0 (0) |
| Arm or leg swelling | 1 (0) | 1 (0) |
| Muscle spasms | 1 (0) | 1 (0) |
| Confusion | 1 (0) | 1 (0) |
| Dyspnea | 1 (0) | 0 (0) |
| Nosebleeds | 0 (0) | 0 (0) |
| Ageusia | 0 (0) | 0 (0) |
| Anosmia | 0 (0) | 0 (0) |
| Conjunctivitis | 0 (0) | 0 (0) |
| Vomiting | 0 (0) | 0 (0) |
| Facial drooping/<br>weakness | 0 (0) | 0 (0) |
| Loss of consciousness | 0 (0) | 0 (0) |
| Seizure | 0 (0) | 0 (0) |

**Figure 2.**
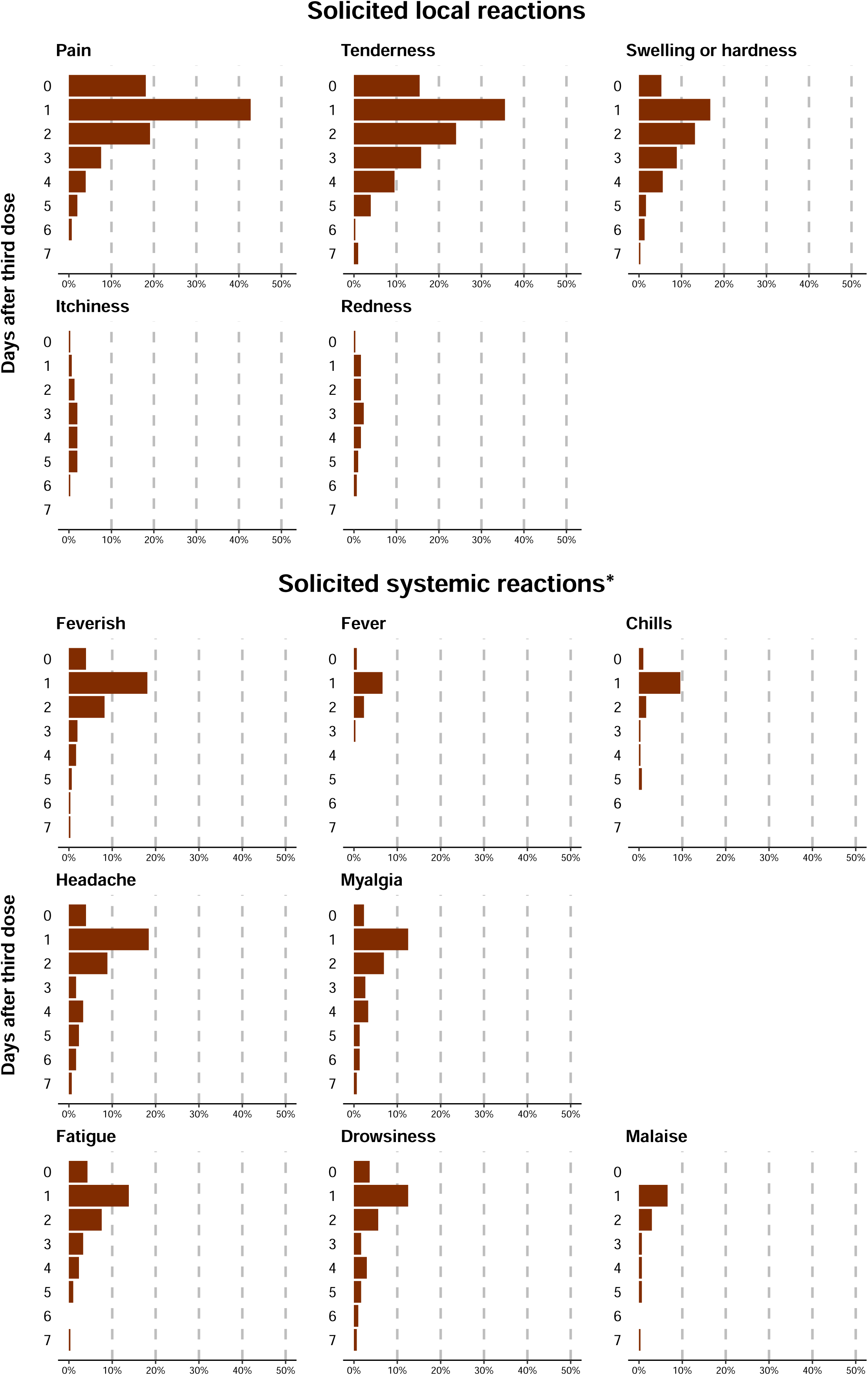
Solicited local and systemic reactions during the 7 days after third dose BNT162b2 vaccination in adults who previously received two doses of inactivated COVID-19 vaccine. *For solicited systemic reactions, only feverish/fever ≥ 38.0°C/ chills, and those reported in at least 10% of participants are shown.

## DISCUSSION

Work by our research group and others indicates that two doses of inactivated COVID-19 vaccine confer moderate increases in antibody levels at one month after the second dose [2, 3]. Antibody levels then gradually decline, but can be boosted by receipt of third doses. Zeng et al. reported that a third dose of inactivated vaccine given to healthy young adults at 8 months after the second dose of the same vaccine boosted neutralizing antibodies against the ancestral strain by about 21-fold [5]. Here, we show that a third dose of mRNA vaccine (BNT162b2) after two doses of inactivated virus vaccine (mostly CoronaVac), all originally formulated against the ancestral virus, boosted PRNT_50_ titers against the ancestral strain by a factor of at least 27-fold. Similarly, Clemens et al. showed that a third dose of mRNA or adenoviral vectored vaccines induced greater rises in anti-Spike binding antibodies compared to a third dose of inactivated vaccine among CoronaVac recipients [9]. Both suggest a substantial improvement of heterologous boosting using mRNA vaccine over homologous boosting using inactivated vaccine against the ancestral virus. We also show that a third dose of BNT162b2 conferred substantial rises in PRNT_50_ titers against the Omicron variant. While there is still a lack of consensus on the antibody threshold for protection [18-21], the PRNT_50_ titers against Omicron at Day 28 in our study are higher than the PRNT_50_ titers against ancestral virus after two doses of CoronaVac where the geometric mean PRNT_50_ titer was estimated to be 27 [2]. Given that two doses of inactivated vaccine were estimated to provide at least 50% protection against infection with the ancestral virus [22-24], the antibody levels against Omicron estimated in our study could correspond to a moderate degree of effectiveness in protection against Omicron.

Most of the participants in our study were older adults, and their antibody levels against Omicron were at a very low level at Day 0 prior to the third dose, consistent with other recent studies [8,9]. Almost one-third of participants had underlying medical conditions (Table 1). In our separate initial investigation which included sera from individuals with various intervals (51-234 days) between second and third dose [11] and some of whom were selected for third dose vaccination due to low (<60%) post-second dose serum surrogate neutralizing antibodies [25], we showed that administering BNT162b2 about 3 months after two doses of CoronaVac reported a GMT of 59 against Omicron compared to 305 against the ancestral virus after the boost [11]. Costa Clemens et al. showed that heterologous boosting with third dose of mRNA or adenoviral vectored vaccines induced greater rise in neutralising antibodies against both Omicron and Delta variants than homologous boosting with inactivated vaccines [9]. Together, these studies suggest the substantial boost in neutralizing antibody titers against both the ancestral virus and Omicron variant by a heterologous third dose of mRNA vaccination will clearly improve protection against the Omicron strain (and potentially other variant strains).

Earlier studies have investigated post-vaccination reactions after two or three doses of inactivated [5, 23, 26] or mRNA vaccination [27,28]. These studies suggest fatigue, myalgia and chill were common both after receipt of inactivated [23] or mRNA vaccines [27]; nausea was common after inactivated vaccination [23]; and fever and headache after mRNA vaccination [27]. Similar to reactions after mRNA vaccination, feverishness and headache were commonly reported in our study while gastrointestinal symptoms such as nausea and diarrhoea were less commonly reported. No hospitalization was reported within the month after vaccination in our study, although our sample size would not have been large enough to detect rare events.

Our study had several limitations. We have only performed PRNT against ancestral strain and Omicron variant up to a dilution of 1:320. However, more than half of our study participants were positive for antibodies to the ancestral strain at this dilution indicating antibody titers ≥320, and we were therefore not able to determine exactly the GMT against ancestral strain after the third dose of vaccination but only confirm that it was likely ≥320. Similarly, Day 0 samples were typically negative even at the starting dilution of 1:10, limiting our assessment of pre-vaccination GMTs. Nevertheless, we were able to conclude that third dose of mRNA vaccination would provide substantial benefits against both ancestral and Omicron strains. Our results on immunogenicity may be subjected to selection bias including volunteer bias, since in Hong Kong older adults are more inclined to receive the inactivated vaccines and younger adults to mRNA vaccines [29]. Our results on reactogenicity may be subjected to information bias as this was an open-label trial and it is recognized that there could be more post-vaccination reactions after mRNA vaccination compared to inactivated vaccination [30].

## Data Availability

Anonymized raw data and R syntax to reproduce all the analyses, figures and tables in the published article will be made available after publication.

## Acknowledgements

We gratefully acknowledge colleagues including Eileen Yu, Teresa So and Zacary Chai for technical support in preparing and conducting this study; Anson Ho for setting up the database; Julie Au and Lilly Wang for administrative support; Hetti Cheung, Victoria Wong, Bobo Yeung at HKU Health System; Cindy Man and other colleagues at the HKU Community Vaccination Centres at Gleneagles Hospital; and all the study participants for facilitating the study.

## Author contributions

All authors meet the ICMJE criteria for authorship. Each author’s contributions to the paper are listed below according to the CRediT model:

Conceptualization: NHLL, GML, BJC

Methodology: NHLL, SMSC

Formal analysis: NHLL, MM-S

Investigation: NYMA, YYN, LLHL, KCKC, JKCL, YWYL, LCHT, SC, KKHK

Funding acquisition: BJC

Project administration: NHLL, SMSC, JSMP, BJC

Supervision: NHLL, SMSC, DKMI, LLMP, GML, JSMP, BJC

Writing – original draft: NHLL, MM-S, JSMP, BJC

Writing – review & editing: NHLL, SMSC, MM-S, NYMA, YYN, LLHL, KCKC, JKCL, YWYL, LCHT, SC, KKHK, DKMI, LLMP, GML, JSMP, BJC.

## FIGURE LEGENDS

**Figure S1.**
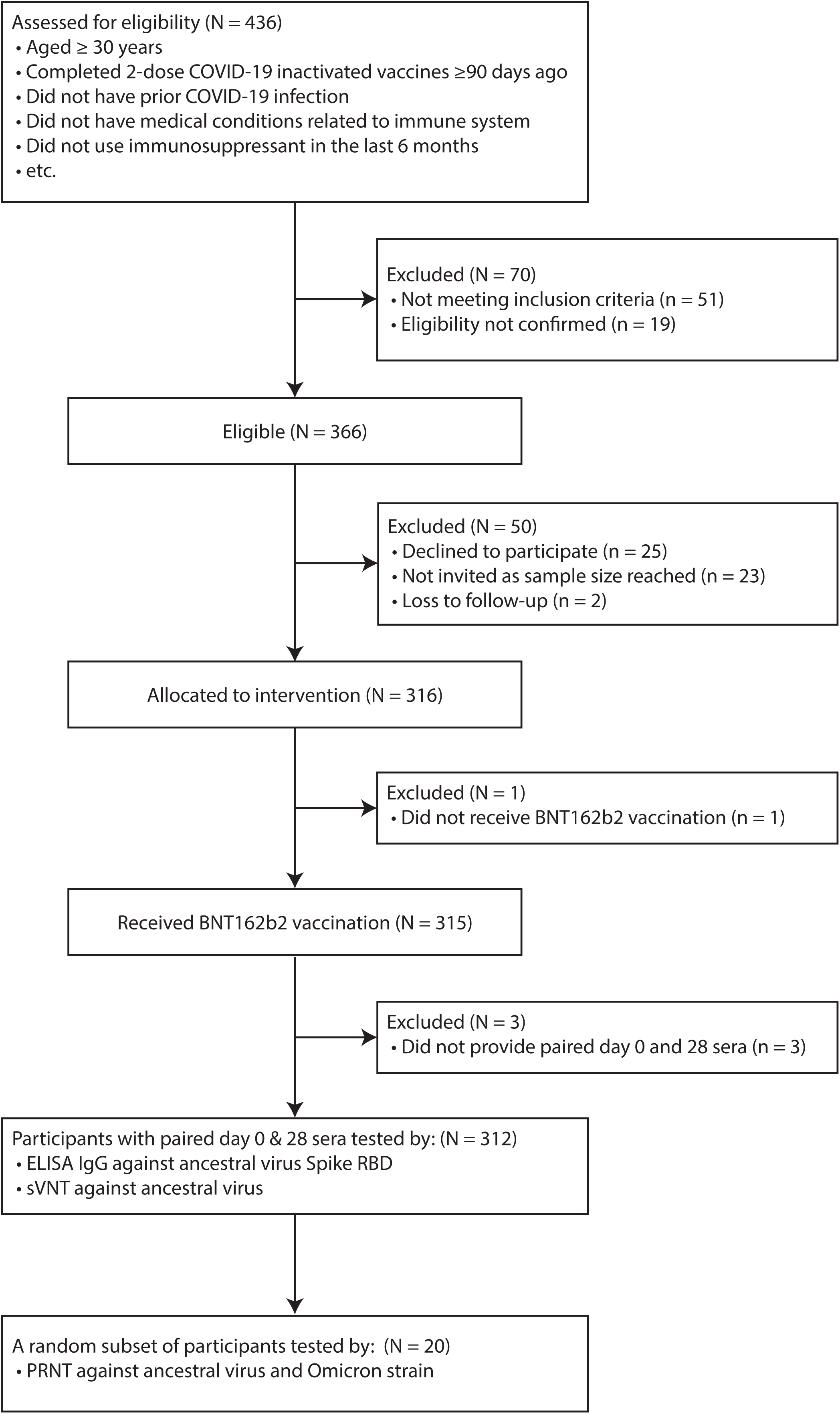
Flow chart for this single-arm, open-label clinical trial of BNT162b2 provided as a third vaccine dose in adults who previously received two doses of inactivated COVID-19 vaccine.

**Figure S2.**
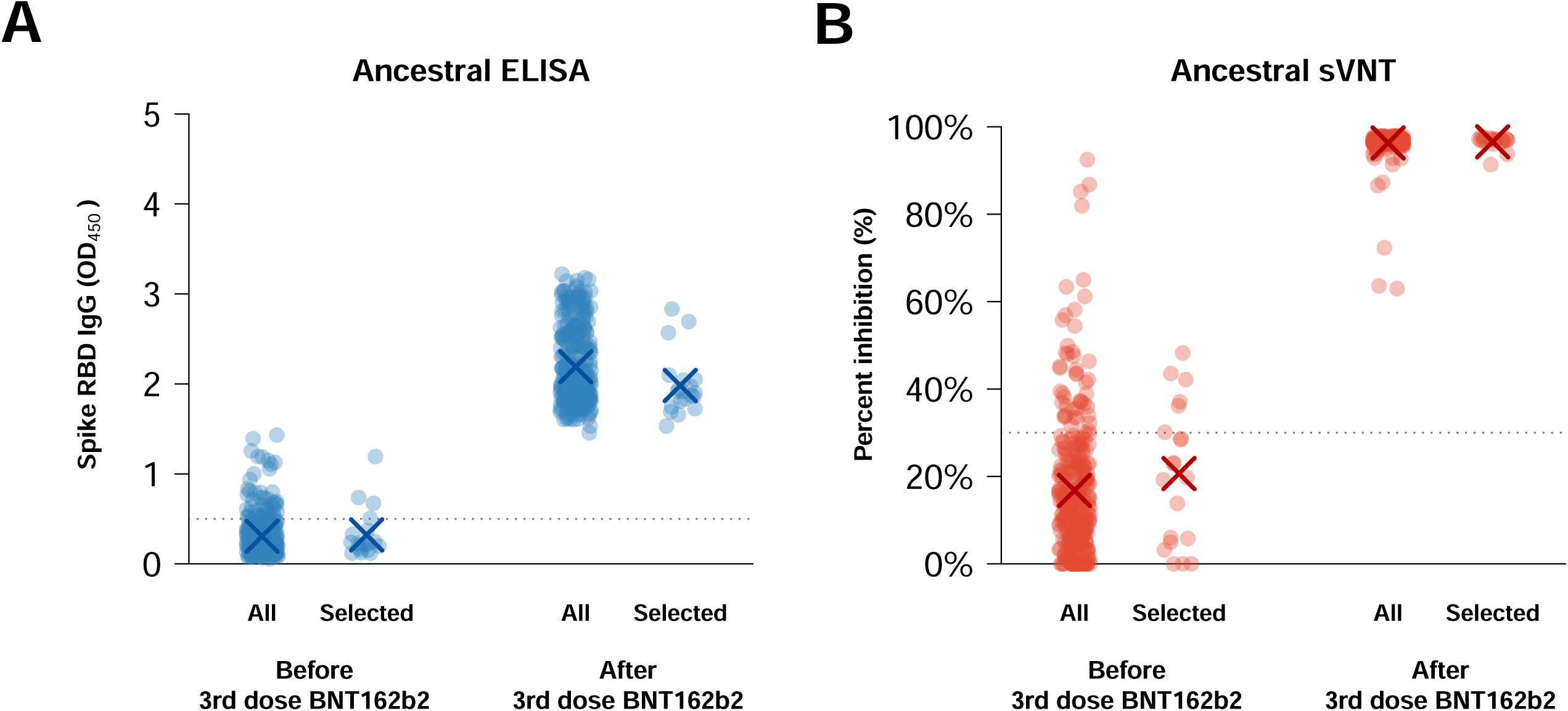
ELISA and sVNT antibody levels in 20 participants randomly selected to be tested by PRNT compared to all vaccinated participants with paired Day 0 and Day 28 sera collected. **(A)** ELISA for serum IgG against SARS-CoV-2 Spike receptor binding domain (RBD) of ancestral strain. **(B)** Surrogate virus neutralization test (sVNT) against ancestral strain. X indicates the mean level. All: all vaccinated participants with paired Day 0/ Day 28 sera collected (N = 312); Selected: participants randomly selected to be tested by PRNT (N = 20).

**Figure S3.**
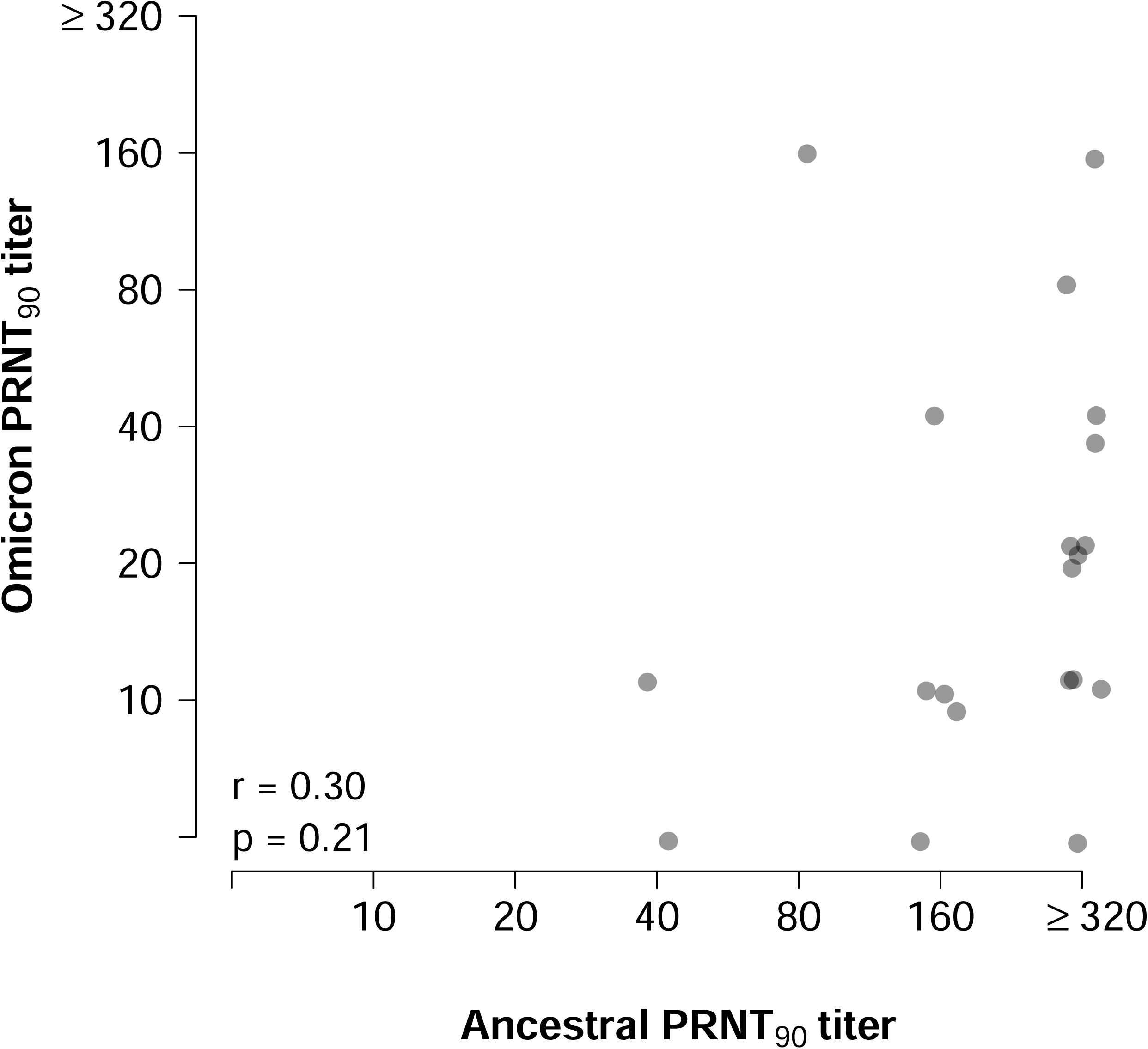
Correlation of PRNT_90_ titers measured at Day 28 against ancestral strain and the Omicron variant in 20 participants randomly selected for performing PRNT. PRNT titers are measured based on two-fold dilutions and plotted here within an interval, or <10 (plotted on axis) or ≥320 (plotted above 320). r: Spearman’s rank correlation coefficient; p: p-value.

**Supplementary Table 1.**
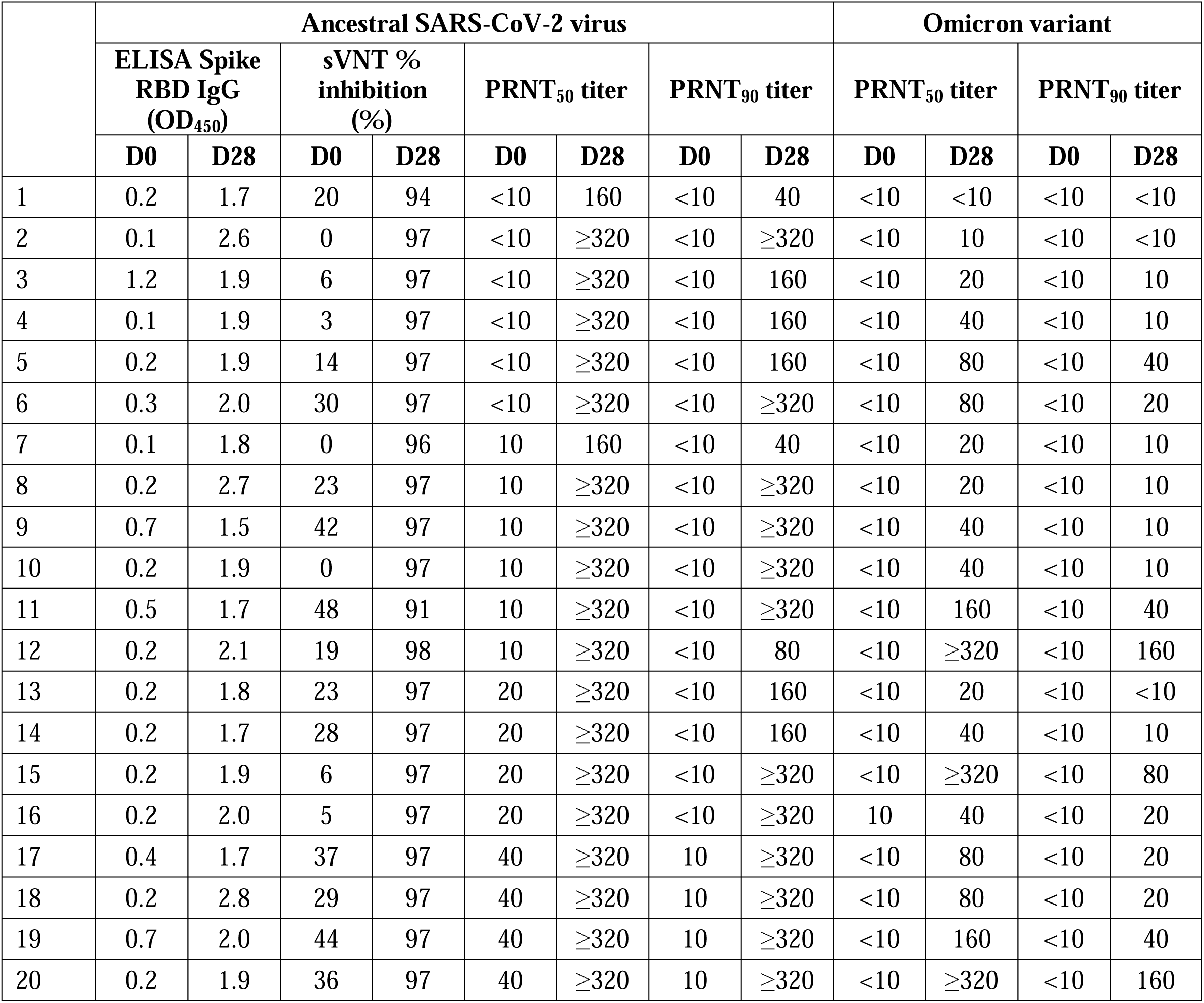
Antibody levels measured by ELISA, sVNT and PRNT against ancestral SARS-CoV-2 virus and the Omicron variant at Day 0 and 28 after third-dose BNT162b2 vaccination, in 20 randomly selected participants who previously received two doses of inactivated COVID-19 vaccine. D0: Day 0 (baseline); D28: Day 28 (post-vaccination); ELISA: enzyme-linked immunosorbent assay; sVNT: surrogate virus neutralisation test; PRNT: plaque reduction neutralisation test; PRNT_50_: 50% plaque reduction neutralization titer; PRNT_90_: 90% plaque reduction neutralization titer

